# MIAMI-AD (Methylation in Aging and Methylation in AD): an integrative knowledgebase that facilitates explorations of DNA methylation across sex, aging, and Alzheimer’s disease

**DOI:** 10.1101/2023.12.04.23299412

**Authors:** David Lukacsovich, Deirdre O’Shea, Hanchen Huang, Wei Zhang, Juan I. Young, X. Steven Chen, Sven-Thorsten Dietrich, Brian Kunkle, Eden R. Martin, Lily Wang

## Abstract

Alzheimer’s disease (AD) is a common neurodegenerative disorder with a significant impact on aging populations. DNA methylation (DNAm) alterations have been implicated in both the aging processes and the development of AD. Given that AD affects more women than men, it is also important to explore DNAm changes that occur specifically in each sex. We created MIAMI-AD, a comprehensive knowledge base containing manually curated summary statistics from 97 published tables in 37 studies, all of which included at least 100 participants. MIAMI-AD enables easy browsing, querying, and downloading DNAm associations at multiple levels – at individual CpG, gene, genomic regions, or genome-wide, in one or multiple studies. Moreover, it also offers tools to perform integrative analyses, such as comparing DNAm associations across different phenotypes or tissues, as well as interactive visualizations. Using several use case examples, we demonstrated that MIAMI-AD facilitates our understanding of age-associated CpGs in AD and the sex-specific roles of DNAm in AD. This open-access resource is freely available to the research community, and all the underlying data can be downloaded. MIAMI-AD (https://miami-ad.org/) facilitates integrative explorations to better understand the interplay between DNAm across aging, sex, and AD.

## BACKGROUND

Alzheimer’s disease (AD) is the most common neurodegenerative disorder, with late-onset AD affecting approximately 1 in 9 people over the age of 65 in the US [1]. AD has become a major public health concern and one of the most financially costly diseases [2]. Among the many risk factors for AD, advanced aging has the strongest impact, with the percentage of people affected by AD increasing from 5% for 65-74-year-olds, 14% in 75–84-year-olds, to 35% in those over 85 [3, 4]. Female sex is another important risk factor for AD [5–9], with almost two-thirds of AD patients in the U.S. being women [10], who also experience more rapid cognitive and functional decline after diagnosis [11, 12].

DNA methylation (DNAm) is an epigenetic mechanism that modifies gene expression without altering the underlying DNA sequence [13]. Importantly, changes in DNAm have been implicated in both aging and AD [14–22]. DNAm changes throughout the lifetime; in particular, as people age, DNAm tends to decrease at intergenic regions but increases at many promoter-associated CpG island regions [17]. Remarkably, age-associated DNAm changes have been observed at many loci [14–16, 23, 24]. Recently, a number of DNAm-based epigenetic clocks have been developed as markers of biological aging. Age acceleration, the difference between estimated epigenetic age and chronological age (and its alternative definitions) [25], has been proposed to assess functional decline in a person during aging and to predict mortality [26].

In addition to aging, we and others have also shown that DNAm is integrally involved in AD [19, 27–33]. Notably, two independent epigenome-wide association studies (EWAS) [19, 31] identified and replicated a number of loci robustly associated with AD neuropathology in the brain. Furthermore, elevated DNAm in the *HOXA* gene cluster was shown to be robustly associated with AD neuropathology in three large cohorts [32]. More recently, we prioritized methylation differences that were consistently associated with AD neuropathology in multiple EWAS cohorts using brain tissues [27]. Encouragingly, many recent studies demonstrated that DNAm differences could also be detected in blood samples of AD subjects [34–39]. Our analysis of two large clinical AD datasets (ADNI [40] and AIBL[41]) revealed many blood DNAm differences consistently associated with AD diagnosis in both cohorts [30]. Moreover, our follow-up sex-specific analysis revealed that many DNAm differences in AD are distinct in men and women [29].

However, currently, our understanding of how changes in DNAm during aging contribute to AD, as well as how DNAm influences aging and AD in a sex-specific manner, is still very limited. As DNAm in the blood is relatively stable and can be detected easily [42], an integrative view of DNAm across sex, aging, and AD in both the brain and blood could not only provide new biological insights but also facilitate the development of reliable, minimally invasive, and inexpensive biomarkers for the diagnosis and prognosis of AD.

Although there are existing databases that provide information on DNAm in the context of aging (e.g., GenAge [43]) and AD (e.g., EWAS Atlas [44] and EWAS Catalog [45]), they do not enable integrative analyses of aging, AD, and sex. To fill this gap, we systematically collated results from recent sex-combined and sex-specific (i.e., sex-stratified) studies in aging and AD and developed a novel integrative database of DNAm across sex, aging, and AD, herein referred to as the MIAMI-AD database. The primary purpose of this database is to provide researchers without prior programming background with an easily accessible resource to search and compare CpGs, regions, and/or genes of interest and benefit from the most recent scientific research. Detailed summary statistics and associated visualizations for both sex-combined and sex-specific analyses of aging and AD are included so that researchers can compare the effects of DNAm without the need to retrieve the original articles. As illustrated by the use-case examples described below in this manuscript, the MIAMI-AD database enables new integrative analyses and biological insights into the role of aging, sex, and DNAm in AD. Moreover, access to all associated summary-level data is freely available to the research community, making MIAMI-AD a valuable resource for biomarker research in aging and AD research.

## CONSTRUCTION AND CONTENT

### Selection of studies and datasets

We performed a comprehensive search for relevant publications using PubMed and Google Scholar, with specific keywords “blood DNA methylation”, “Alzheimer’s disease”, “dementia”, “aging”, “sex-specific associations”, and “sex-stratified”. To be included in MIAMI-AD, the published studies must meet three main criteria: (1) having more than 100 total samples from human subjects; (2) conducting a genome-wide study of more than 100k CpGs; and (3) utilizing Illumina 450k or EPIC arrays. For each of these selected studies, we included as many CpGs as possible, even those that did not reach the statistical significance threshold. These nonsignificant CpGs could be valuable for future meta-analyses or pathway analyses. We included studies that utilized brain or blood DNAm data. Supplementary Table 1 contains detailed information for all the studies included in the MIAMI-AD database.

### Web interface and implementation

MIAMI-AD (https://miami-ad.org/) is a web application developed using the Shiny platform [46–48]. All underlying data analysis and management were conducted using R statistical software (https://www.R-project.org/). The web interface is organized into four main sections, each accessible via a tab at the top. Additionally, a left-side panel allows users to select search parameters. For a quick start, each section features a guided tour built using the cicerone R package, which provides a walkthrough of the specific options available in that section.

There are four main sections: Genome-wide Query, Gene Query, CpG Query, and Epigenetic Clock Query. Each of these sections is further divided into three subsections, accessible via subtabs:

1. **Datasets**: Here, users can select the studies or epigenetic clocks they wish to examine. The other subtabs remain hidden until a selection is made in this section.
2. **Display Data**: This section shows tables of summary statistics tailored to the chosen datasets and relevant parameters from the main tabs. Users also have the option to download these tables as Excel files.
3. **Display Plots**: Visual representations of the data can be found here, including Manhattan plots via the R package miamiplot, Venn diagrams via the R package ggvenn, forest plots via the R package meta, and genomic annotations via R package Gviz [49], annotation information from the UCSC track hub https://genome.ucsc.edu/cgi-bin/hgHubConnect, and computed chromatin states from the NIH Roadmap Epigenomics project [50]. The source code for MIAMI-AD can be accessed at https://github.com/TransBioInfoLab/MIAMI.AD.

### Data access and protection of private information

MIAMI-AD offers valuable information on summary statistics of genes, regions, and CpGs related to DNAm differences in aging, AD, and sex. To ensure easy access to the database, we have incorporated “Download Tables” links within each query tool, available under the “Display Data” tab (Figure 1). Researchers can conveniently retrieve their search results using these links. Furthermore, for comprehensive access to the summary statistics of all CpGs in each study, a “Download” tab is provided in the main menu (Figure 1).

**Figure 1.**
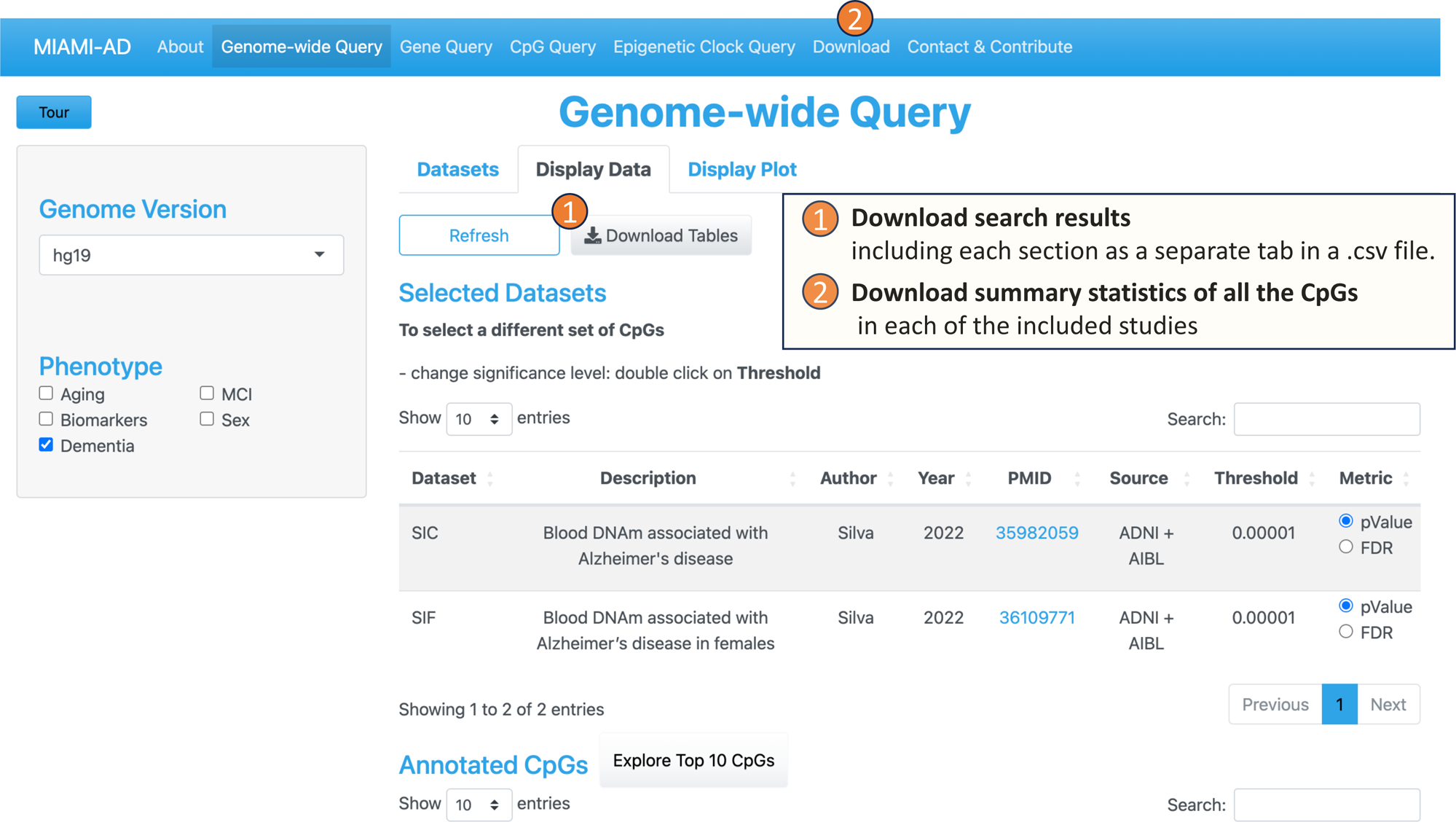
Users can download search results, as well as summary statistics for all the CpGs in each study.

However, it is important to note that MIAMI-AD does not support the access or download of the original datasets used to generate the summary statistics and figures. This includes both phenotypic and epigenomic data. Our focus is solely on providing summary statistics and visualizations based on groups of subjects, without any identifying information on individual study participants. This approach ensures the protection of subjects’ privacy and data confidentiality.

### Technical validation of the database

To ensure the accuracy of the MAIMI-AD database, three coauthors (HH, WZ, and DOS) performed independent verifications. They cross-checked the content of the database against the supplementary tables from the source publications by randomly sampling a list of CpGs, genes, and DMRs in each supplementary table/dataset.

## UTILITY AND DISCUSSION

MIAMI-AD facilitates in-depth exploration of DNA methylation in the brain and the blood at various levels: CpGs, genes, regions, or on a genome-wide scale, specifically related to aging and AD, and across different sexes. The database currently contains both statistically significant and nonsignificant genome-wide summary statistics, as well as epigenetic clock information from 97 main, supplementary, and externally referenced tables recently published in a total of 37 studies (Supplementary Table 1), with a minimum of 100 independent subjects in each study. The platform offers four distinct types of queries to meet different user needs:

1. **The Genome-wide Query tool** is designed to allow users to select CpGs across all chromosomes based on a significance threshold, making it valuable for comparing association results from one or more studies. Figure 2 illustrates the workflow of the Genome-wide Query. First, the user can choose one or more phenotypes of interest from options such as “Aging,” “Biomarkers,” “Dementia,” “MCI,” or “Sex,” available in the left panel. On the right panel, under the “Dataset” tab, a table displays the studies and datasets associated with the selected phenotypes. Here, we refer to “studies” as research publications and “datasets” as the resulting summary statistics from these studies.

**Figure 2.**
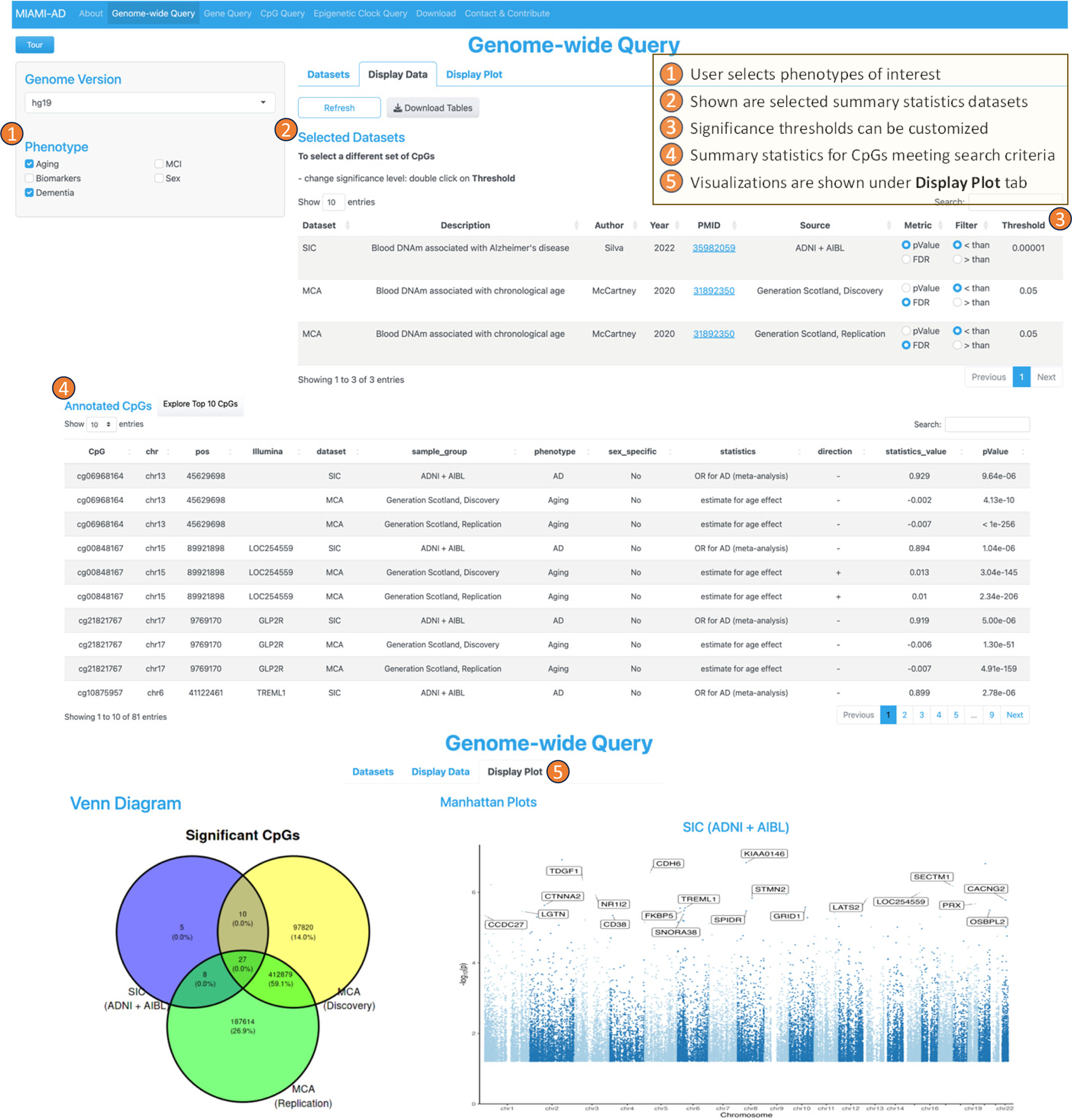
Workflow of the Genome-wide Query tool. In the second step, the user can select datasets of interest by checking the boxes in the rightmost column. Upon doing so, the “Display Data” tab presents the CpGs that meet the search criteria. It includes annotations on location and associated genes, along with summary statistics such as the direction of the association, test statistic values, raw *P* values, and multiple comparison-adjusted *P* values. Additionally, the “Display Plot” tab generates visualizations, such as Venn diagrams to illustrate the numbers of overlapping and unique CpGs across multiple studies, as well as Manhattan plots for genome-wide representation of the data.
2. **The Gene Query tool** is designed to provide detailed information on DNAm differences within a specific gene or genomic region. The interface for this tool is similar to the Genome-wide Query Tool, except that on the left panel, the user additionally specifies the name of the gene (or region) they want to explore and selects the desired annotation tracks such as UCSC gene, ENSEMBL genes, ENSEMBL transcripts, chromatin states, and CpG islands (Figure 3).

**Figure 3.**
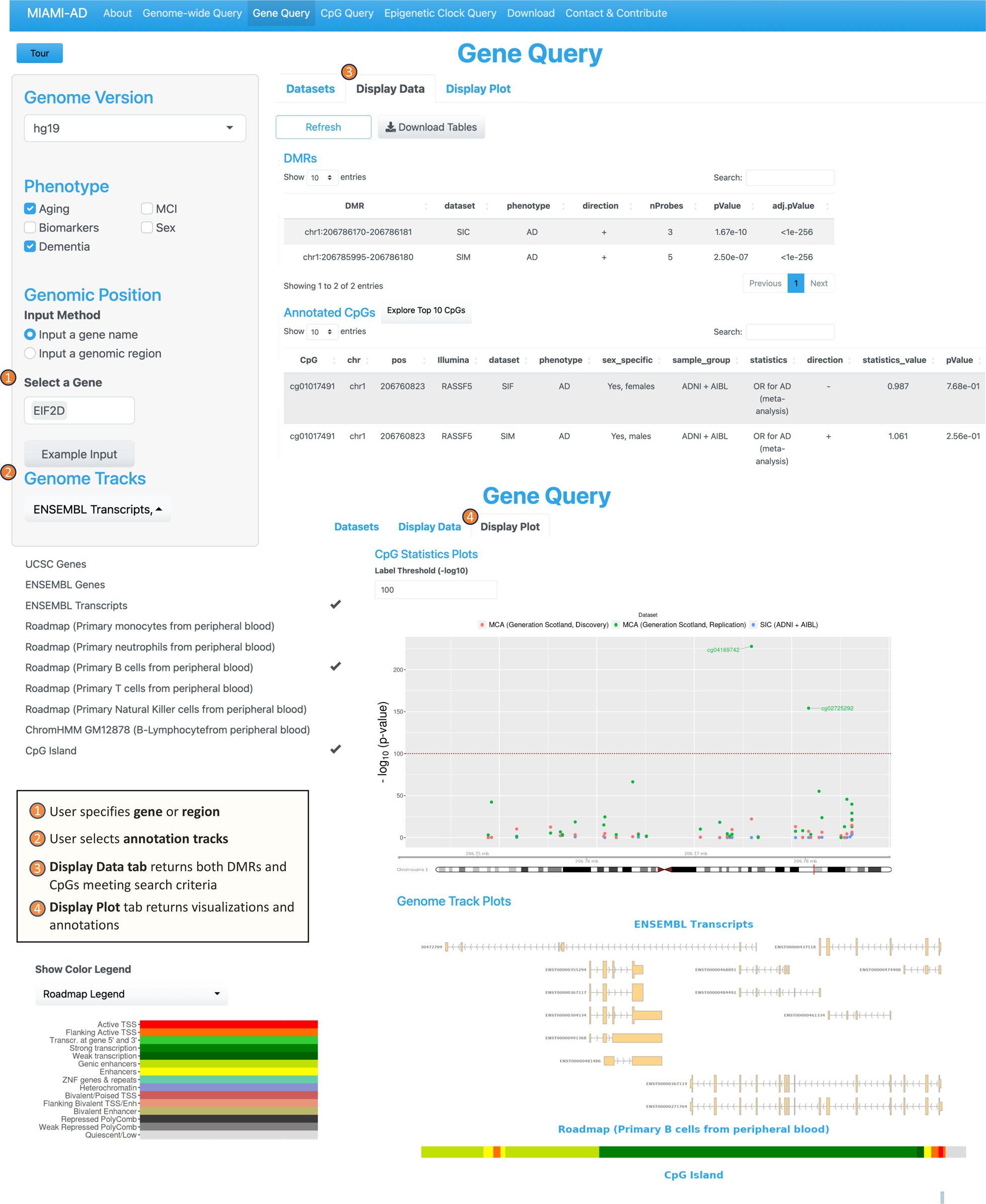
Workflow of the Gene Query tool. Upon completing the inputs, the “Display Data” tab on the right panel presents a summary of statistics for CpGs located within a specified range of ± 2 kb around the gene of interest. This summary includes information on the direction of association and the values of the statistical measures (e.g., odds ratio, t-statistic, and the corresponding *P* values). The “Display Plot” tab generates visualizations, including a mini-Manhattan plot of the CpGs found within the gene, along with the selected annotation tracks such as CpG island, gene transcripts, and computed chromatin states. These plots offer a clear and concise way to interpret the data and understand the relationships between DNAm and the phenotype in the gene or region of interest.
3. **The CpG Query tool** offers information about specific CpGs of interest (Figure 4). To use this tool, the user begins by inputting the desired phenotypes on the left panel and providing a list of CpGs they want to explore. In the right panel, the user selects the relevant studies from the available “Datasets.”

**Figure 4.**
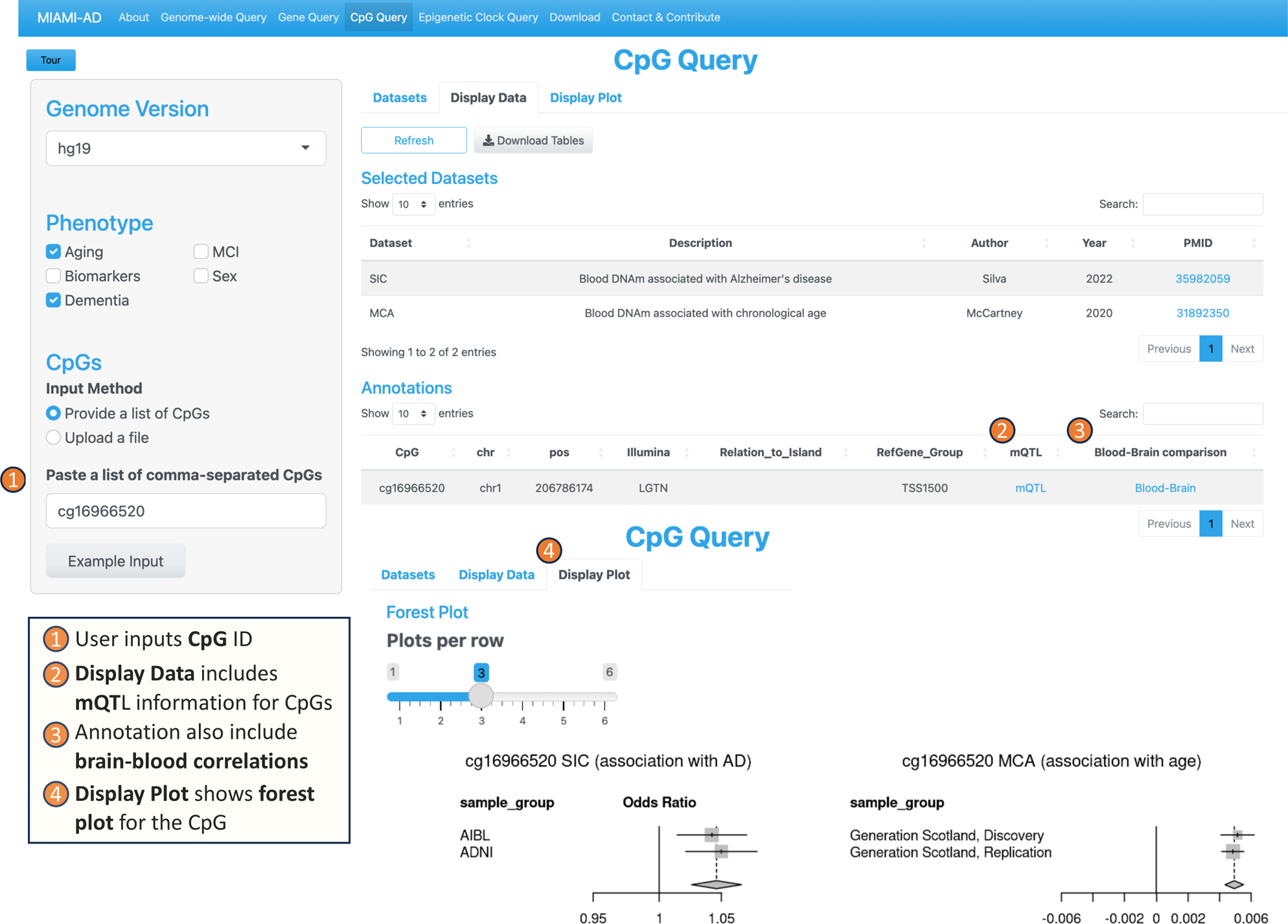
Workflow of the CpG Query tool In the “Display Data” tab, the tool provides not only summary statistics for the selected CpGs but also valuable annotations for each CpG. These annotations include CpG location, genes associated with the CpG, and mQTL (methylation quantitative trait loci) information obtained from a recent large-scale meta-analysis [51]. Additionally, the tool includes the correlation of blood DNAm with brain DNAm at the particular CpG [52], offering insights into potential cross-tissue relationships. Furthermore, MIAMI-AD searches among 14 recently published epigenetic clocks and identifies matches if a selected CpG is a component of any epigenetic clock. Under the “Display Plot” tab, the tool generates forest plots to illustrate the association of the selected CpGs with the specified phenotypes across different datasets. This visualization aids in understanding the relationship between DNAm at CpG sites and the various phenotypes of interest.
4. **The Epigenetic Clock Query tool** serves the purpose of comparing with and selecting CpGs that have been utilized in constructing blood-based epigenetic clocks (Figure 5). This tool incorporates a total of 14 epigenetic clocks, which include the widely described pan-tissue clock by Horvath (2013) [53], as well as the recently developed DunedinPACE clock, which is associated with the risk of dementia onset [54], among others. Including epigenetic clock CpGs allows researchers to perform comparative analyses between EWAS-derived CpGs and those from epigenetic clocks, potentially revealing unique or shared pathways in the aging process. Additionally, given the popularity and widespread use of epigenetic clocks in aging research, their inclusion can attract a broader user base to the platform, making it a go-to resource for epigenetic researchers.

**Figure 5.**
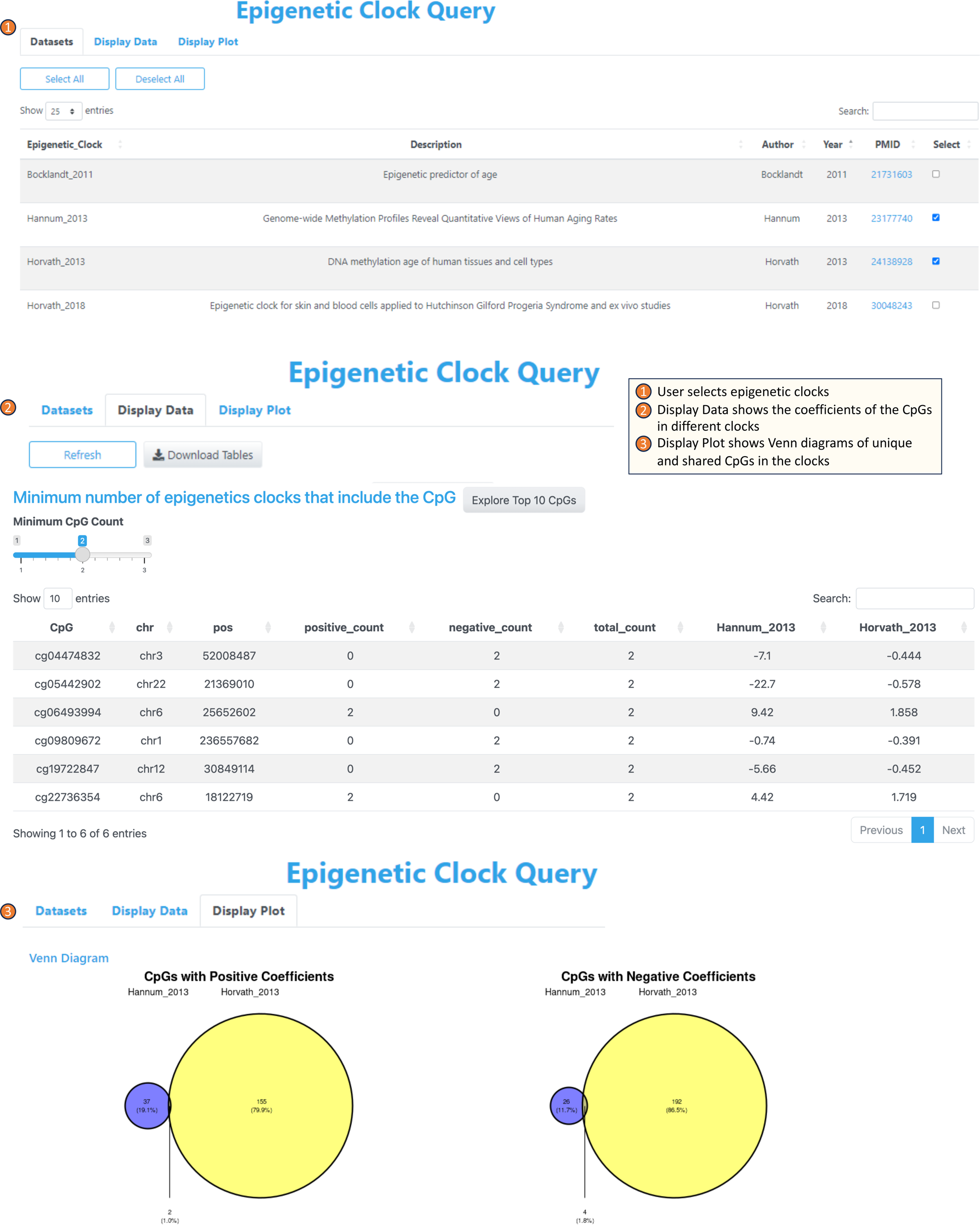
Workflow of the Epigenetic Clock Query tool. To use the Epigenetic Clock tool, users start by choosing the specific epigenetic clocks of interest from the “Dataset” category. Subsequently, upon navigating to the “Display Data” tab, the tool provides a list of the CpGs that are incorporated into one or more of the selected clocks, accompanied by their respective coefficients within each clock. Moving on to the “Display Plot” section, users can view the number of CpGs shared across multiple clocks, as well as those unique to individual clocks, as visualized by Venn diagrams.

### Case study 1: Using Genome-wide Query tool to identify DNA methylation differences in both aging and AD

Aging and AD are intertwined, while molecular processes in aging can promote AD, AD pathology can also influence aging processes [55]. Although a common view is that DNAm changes that occur during aging are accentuated in AD [56, 57], recent studies by Berger and colleagues suggested that there are also epigenetic changes that normally occur in aging, perhaps protective, that are disrupted in AD [58, 59]. McCartney et al. (2020) performed an EWAS of chronological aging using the Generation Scotland datasets and revealed pervasive changes in the epigenome during aging [60]. To study the role of age-associated CpGs in AD, we compared the CpGs significantly associated with chronological age with those significantly associated with AD using MIAMI-AD. Specifically, we compared the CpGs with a 5% false discovery rate (FDR) from both discovery and replication analyses in McCartney et al. (2020) with those in our recent meta-analysis of blood DNAm samples in AD [30], in which we identified 50 significant CpGs that were consistently associated with clinical AD in the ADNI and AIBL studies (i.e., meta-analysis *P* value < 10^−5^).

Using the Genome-wide Query Tool, we selected 27 CpGs that are significant in both aging and AD studies (Figure 2). Next, we obtained Supplementary Table 2 via the “Download Tables” button under “Display Data”, which shows that among these 27 CpGs, two-thirds (18 CpGs) had the same direction of change in their associations with chronological age and AD, while one-third (9 CpGs) had opposite directions of change. In the next section, we explore the role of these CpGs in aging and AD in more detail.

### Case Study 2: Using the CpG Query tool to understand the roles of aging-associated CpGs in AD

One interesting example is cg16966520 located in the promoter of the *EIF2D* gene. Using the CpG Query tool, we obtained forest plots that illustrate DNA methylation changes of cg16966520 in aging and AD. In Figure 4, the forest plot in the lower right corner shows that this CpG is significantly hypermethylated in aging in both the discovery and replication datasets of McCartney et al. (2020) [60]. Moreover, the forest plot on the left shows significant positive associations between DNA methylation at cg16966520 and clinical AD in both the ADNI and AIBL datasets in our previous meta-analysis of clinical AD [30]. Therefore, cg16966520 is an example of an *age-associated CpG with an amplified effect* in AD.

The *EIF2D* gene encodes the eukaryotic translation initiation factor 2D, which plays an important role in protein translation [61]. Previous studies have observed reduced protein synthesis during aging, accompanied by a decrease in ribosome abundance, attenuated activity, and decreased concentrations of predominant initiation and elongation factors [62, 63]. The observed hypermethylation in the promoter region of the *EIF2D* gene is consistent with these findings, particularly the decreased levels of protein synthesis initiation factors. Interestingly, we also observed hypermethylation of this locus in the blood samples of AD subjects [30], suggesting that hypermethylation at cg16966520 became even more prominent in AD subjects. Furthermore, the link to the Blood‒Brain Comparison Database [52] under “Annotations” in the CpG Query tool shows that blood DNAm at cg16966520 is significantly correlated with DNAm in the brain prefrontal cortex (r = 0.618, *P* value = 4.44×10^−9^), entorhinal cortex (r = 0.511, *P* value = 5.40×10^−6^), superior temporal gyrus (r = 0.555, *P* value = 2.43×10^−7^), and cerebellum (r = 0.378, *P* value = 1.16×10^−3^), indicating that this age-associated CpG is a potential biomarker for AD (Supplementary Figure 1).

Another interesting CpG that overlapped between our meta-analysis of AD [30] and the aging EWAS [60] is cg13270055, located on the *CACNG2* gene. Using the CpG Query tool, we obtained forest plots of DNAm changes at cg13270055 in aging and AD, highlighting the divergent trends in changes. Specifically, Supplementary Figure 2 shows that cg13270055 is significantly hypermethylated in aging in both the discovery and replication datasets of McCartney et al. (2020). Interestingly, this CpG is also significantly hypomethylated in AD subjects in both the AIBL and ADNI datasets.

The *CACNG2* gene, which encodes a transmembrane protein that modulates neurotransmission of glutamate receptors, plays important roles in learning, memory, and synaptic plasticity (the formation of new synapses between neurons) [64]. Interestingly, recent studies have linked the *CACNG2* gene to stress-coping mechanisms [65]. It has been observed that the brain expression levels of the *CACNG2* gene were increased in both monkeys and mice after they were exposed to repeated temporal stress conditions and stress cope training sessions [64]. These results are supported by another study that found decreased *CACNG2* gene expression levels in the prefrontal cortex of schizophrenia patients [66]. In AD, consistent with our observed hypomethylation at the *CACNG2* gene body [30], another study also noted decreased *CACNG2* gene expression in the hippocampus of AD patients [67].

These results are consistent with the recent theory that synapse failure is an early feature of AD [68] and that synaptic plasticity promotes cognitive reserve, the ability of the brain to maintain normal cognitive function despite the presence of significant Alzheimer’s disease brain pathology [69, 70]. Therefore, consistent with earlier results of Nativio et al. (2018; 2020) [58, 59], we found that some DNAm changes (e.g., hypermethylation at cg13270055) in aging may also be disrupted or fail to be established in AD. Encouragingly, the link to the Blood‒Brain Comparison Database [52] under “Annotations” within the CpG Query tool shows that blood DNAm at cg13270055 is also significantly correlated with DNAm in the brain cortex regions, including the prefrontal cortex (r = 0.414, *P* value = 2.46×10^−4^), entorhinal cortex (r = 0.497, *P* value = 1.05×10^−5^), and superior temporal gyrus (r = 0.468, *P* value = 2.31×10^−5^), indicating that these CpGs are plausible biomarkers for AD (Supplementary Figure 3).

### Case study 3: Understanding the sex-specific roles of DNA methylation in Alzheimer’s disease

Sex is increasingly recognized as a significant factor contributing to the biological and clinical heterogeneity in AD [5–9]. To study the role of sex-specific DNAm in AD, we used the Genome-wide Query tool to compare DNAm to AD associations of men vs. women in sex-stratified analyses of Silva et al. (2022) [29]. First, to examine whether there was any overlap between significant CpGs in the analysis of female samples and those in male sample analysis, we selected the “SIF” and “SIM” datasets, corresponding to female and male summary statistics datasets in Silva et al. (2022). This input did not return any CpG with a *P* value < 10^−5^ in both SIF and SIM, indicating that the significant CpGs in the analysis of female samples are distinct from those in the analysis of male samples (Supplementary Figure 4A).

To select female-specific CpGs associated with AD, we changed the significance threshold of the Genome-wide Query tool to a *P* value < 10^−5^ for the female dataset (SIF) and a *P* value > 0.05 for the male (SIM) dataset (Supplementary Figure 4B). This search resulted in 21 CpGs, which can be downloaded by selecting the “Download Tables” button below “Display Data” (Supplementary Table 3).

Next, using the CpG Query tool, we examined these sex-specific DNAm in more detail. An interesting CpG is cg03546163 on the *FKBP5* gene. In Supplementary Figure 5, the summary statistics under “Display Data” show that this CpG is significantly hypomethylated in women in both the ADNI and AIBL cohorts (OR_ADNI_ = 0.924, *P*_ADNI_ = 3.02×10^−3^, OR_AIBL_ = 0.896, *P*_AIBL_ = 8.86×10^−5^) but is not significant in men. It is worth noting that prior research has associated genetic variations in *FKBP5* with major depressive disorder [71, 72], a significant risk factor for dementia and with a higher prevalence among women [73].

Notably, our previous sex-combined meta-analysis [30] also identified cg03546163. Here, the sex-specific analysis provided additional insight that the significant DNAm-to-AD association at cg03546163 is predominantly driven by effects in women. This example demonstrates that MIAMI-AD can be applied to significant CpGs identified in sex-combined analysis to better understand their sex-specific roles in AD.

Similarly, we can also use MIAMI-AD to identify male-specific DNAm associated with AD by selecting a *P* value < 10^−5^ for the men dataset (SIM) and a *P* value > 0.05 for the women dataset (SIF) (Supplementary Figure 4C). This search returned 4 CpGs, which we downloaded using the “Download Tables” button in Genome-wide Query (Supplementary Table 4). An interesting male-specific CpG is cg15757041, located in the promoter region of the *C16orf89* gene. Under “Display Data”, the CpG Query tool showed that in men, this CpG was significantly hypomethylated in AD subjects in both the ADNI and AIBL cohorts (OR_ADNI_ = 0.695, *P*_ADNI_ = 1.94×10^−4^; OR_AIBL_ = 0.625, *P*_AIBL_ = 5.14×10^−3^). On the other hand, this CpG is not significant in women. The *C16orf89* gene is mainly expressed in the thyroid and is involved in thyroid function. Our result is consistent with recent studies that linked thyroid dysfunction with AD [74, 75].

### Discussion and future directions

Alzheimer’s disease (AD) remains a significant public health concern as the aging population continues to grow. Here, we introduced the MIAMI-AD database, a new resource that offers an integrated perspective on DNA methylation in aging, sex, and AD. By collating a large number of summary statistics datasets from recent studies, MIAMI-AD provides a comprehensive overview of DNAm changes associated with aging, sex, and AD in brain and blood samples, enabling researchers to explore the intricate relationships between these factors.

The results presented in this manuscript demonstrate the utility and versatility of the MIAMI-AD database. Through case studies, we demonstrated how researchers can leverage the database to uncover novel insights into the roles of DNAm in aging and AD. The Genome-wide Query tool facilitates the identification of shared and distinct DNAm patterns between aging and AD, shedding light on the complex interplay between these processes. Moreover, the gene and CpG Query tools empower researchers to delve into specific genomic regions of interest, allowing for detailed investigations into the functional implications of DNAm changes.

Importantly, MIAMI-AD also recognizes the growing significance of sex-specific associations in AD research. By including sex-stratified analyses, the database revealed distinct DNAm patterns that contribute to the observed heterogeneity in AD between men and women. This enables new insights into sex-specific effects and underscores the need for personalized approaches to understanding AD pathogenesis.

To the best of our knowledge, MIAMI-AD is the first comprehensive knowledgebase focused on an integrative view of DNAm in aging, sex, and AD. There are several alternative, more general databases dedicated to EWAS and aging. For example, both the EWAS Atlas [44] and EWAS Catalog [45] curated a wealth of knowledge including hundreds of human traits across diverse tissues and cell lines. Additionally, the Aging Atlas database [76] curates multiomics data in aging, which includes findings discovered in ChIP-seq, RNA-seq, proteomics, and other high-throughput technologies. However, these sources do not include DNA methylation data. In contrast, MIAMI-AD focuses on presenting and disseminating EWAS results obtained in brain and blood samples in recent dementia research, which is particularly relevant to the development of DNAm-based biomarkers for dementia.

MIAMI-AD incorporates a large number of high-quality associations from recent publications in aging and dementia research. To ensure accuracy, the content of the database was independently validated against original publications by three coauthors, HH, WZ, and DOS (Supplementary Table 5). This open-access resource is freely available to the research community, and all the underlying data can be downloaded. In addition, it also allows users to report any issues encountered and contribute additional published results, which expands the AD knowledgebase and its potential impact.

MIAMI-AD offers a wealth of insights into sex, aging, and AD. Our future efforts include expanding the scope of MIAMI-AD to connect EWAS associations with GWAS associations, along with other types of molecular changes related to AD, such as gene expression, proteins, and metabolites in diverse tissues. MIAMI-AD will be regularly updated to incorporate associations from the latest aging and AD literature.

## CONCLUSIONS

DNA methylation offers great promise in advancing our understanding of dementia. We expect MIAMI-AD to be an invaluable resource that fosters collaboration and knowledge sharing among researchers. With its user-friendly interface, open access to the research community, comprehensive datasets, and potential for expansion, MIAMI-AD will play important roles in enabling new biological insights into the relationship between DNAm, aging, and sex in AD. Moreover, it will help with prioritizing blood biomarkers, which will facilitate the development of innovative diagnostic and therapeutic strategies for this global health challenge.

## Supporting information

All_Supp_Tables

## Data Availability

All datasets analyzed in this study are publicly available and listed in Supplementary Table 1. The scripts for the analysis performed in this study can be accessed at https://github.com/TransBioInfoLab/MIAMI.AD

## DECLARATIONS

### Ethics approval and consent to participate

Not Applicable

### Consent for publication

Not Applicable

### Author Contributions

L.W., D.O. S, D.L., J.I.Y., E.R.M., B.K., X.S. C designed the database and computational analyses. D.L. analyzed the data and created the database. S.D. provided technical assistance and testing of the database. L.W., D.O. S, J.I.Y., E.R. M, B.K., X.S.C. contributed to the interpretation of the results in case studies. H.H., W.Z., and D.O.S. independently validated the content of the database against original publications. L.W. and D.L. wrote the paper, and all authors participated in the review and revision of the manuscript. L.W. conceived the original idea and supervised the project.

## Acknowledgment

We thank the members of the UMIT team and Dean Attali for their technical assistance during the development of the database.

---

**Supplementary Figure 1.**
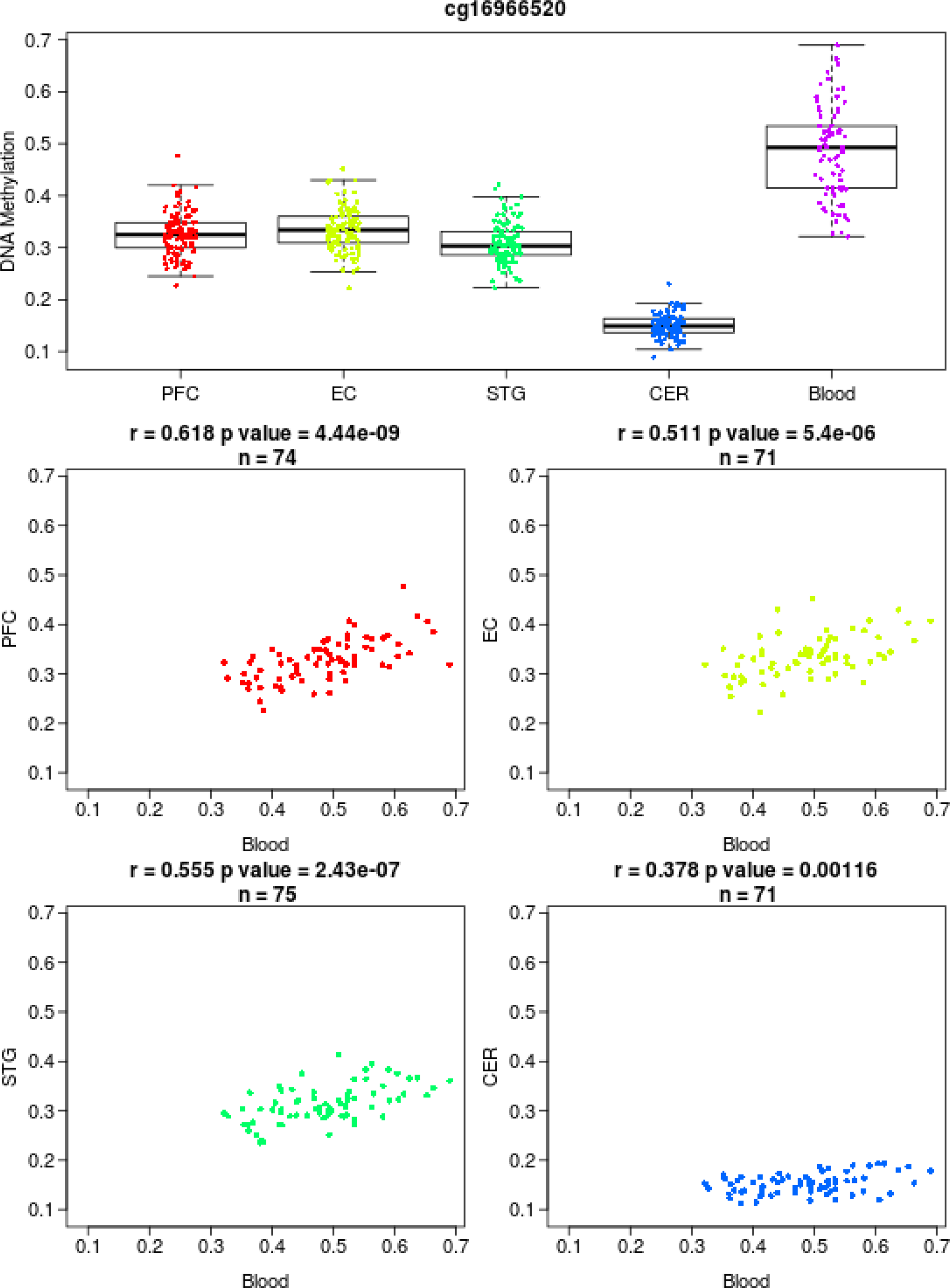
Blood DNA methylation at cg16966520 in the promoter region of the *EIF2D* gene is significantly associated with brain DNA methylation in the prefrontal cortex (PFC), entorhinal cortex (EC), superior temporal gyrus (STC), and cerebellum (CER). These results were obtained from the Blood Brain DNA Methylation Comparison Tool (https://epigenetics.essex.ac.uk/bloodbrain/).

**Supplementary Figure 2.**
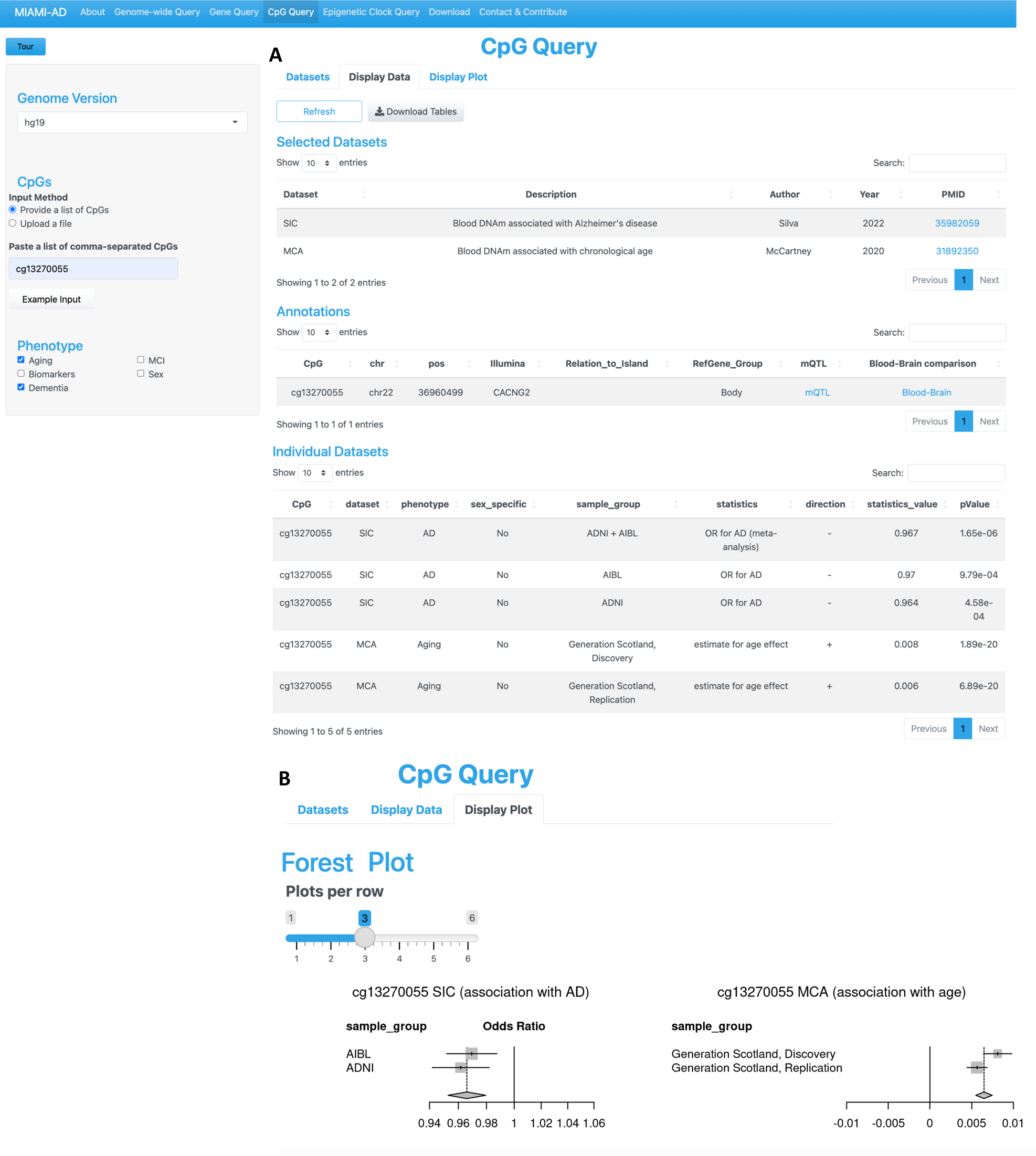
DNA methylation at cg13270055 located on the *CACNG2* gene increases with age (*P*_discovery_ = 1.89 ×10^−20^, *P*_replication_ = 6.89 ×10^−20^), but decreases in the blood samples of Alzheimer’s subjects (*P*_meta-analysis_ = 1.65×10^−6^, *P*_AIBL_ = 9.79×10^−4^, *P*_ADNI_ = 4.58×10^−4^). The summary statistics are shown under **(A)** Display Data tab, and the forest plots are shown under **(B)** Display Plot tab.

**Supplementary Figure 3.**
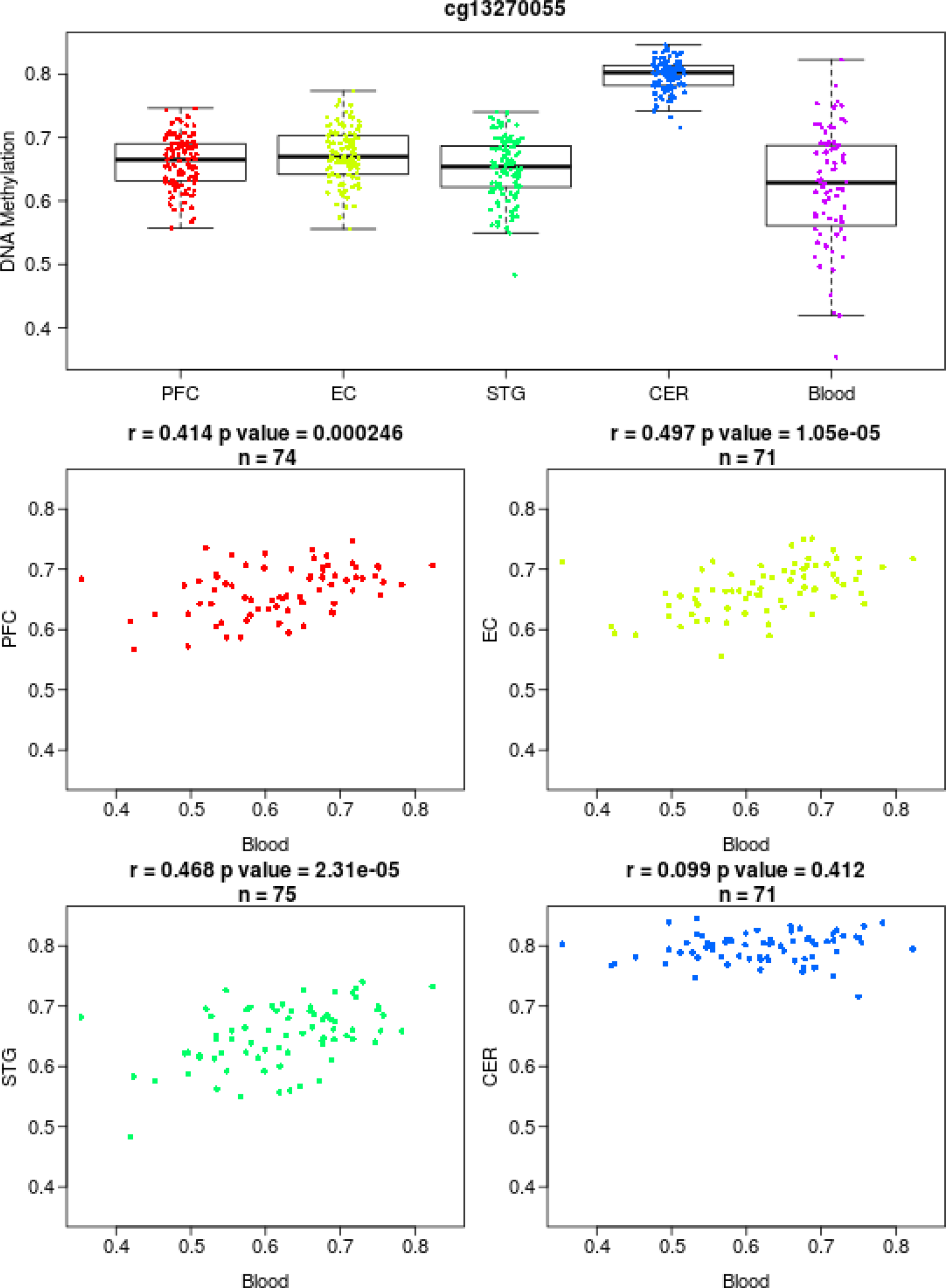
Blood DNA methylation at cg13270055 located on the *CACNG2* gene is significantly associated with brain DNA methylation in the prefrontal cortex (PFC), entorhinal cortex (EC), superior temporal gyrus (STC), and cerebellum (CER). These results were obtained from the Blood Brain DNA Methylation Comparison Tool (https://epigenetics.essex.ac.uk/bloodbrain/).

**Supplementary Figure 4.**
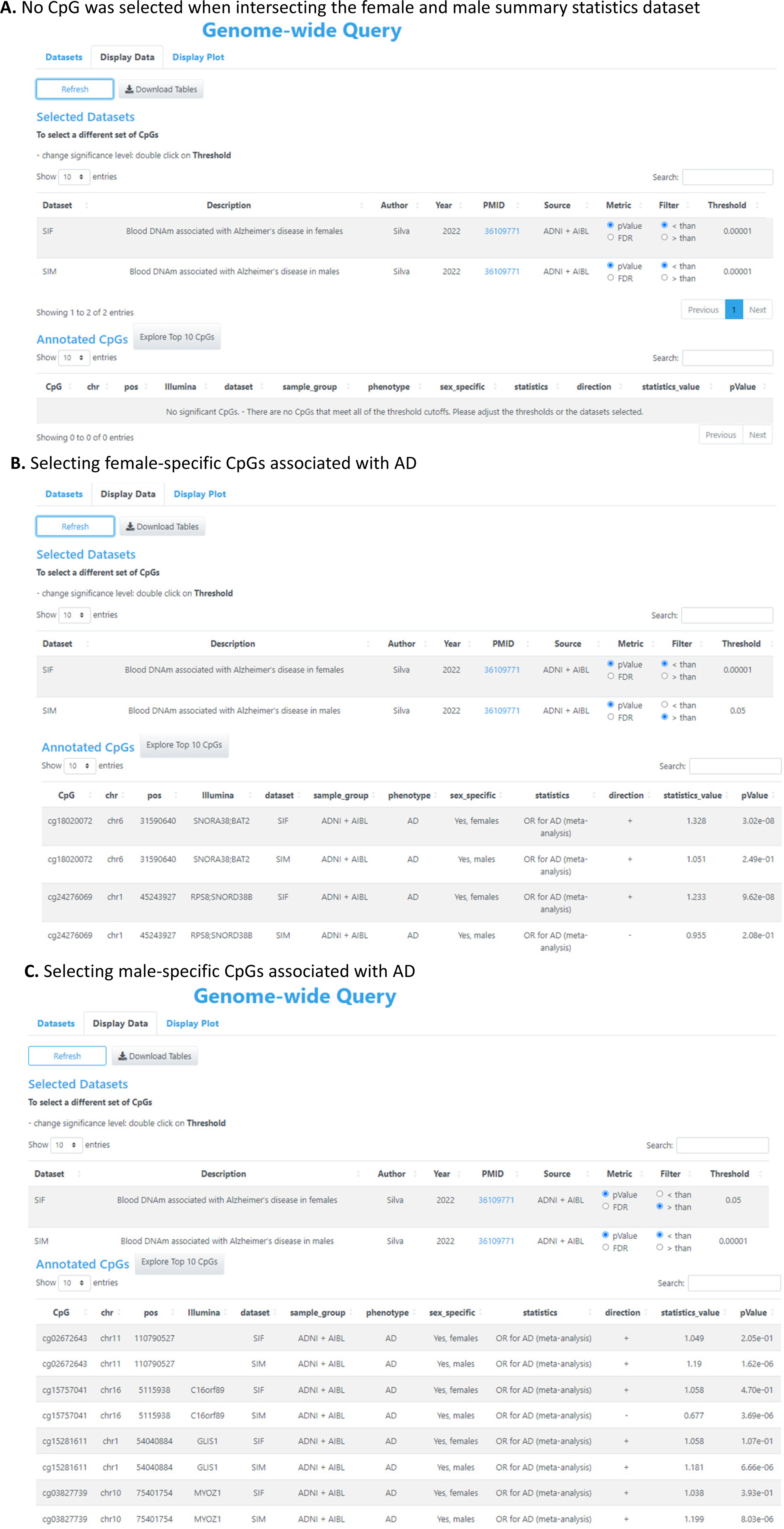
Using Genome-wide Query to perform sex-specific analysis.

**Supplementary Figure 5.**
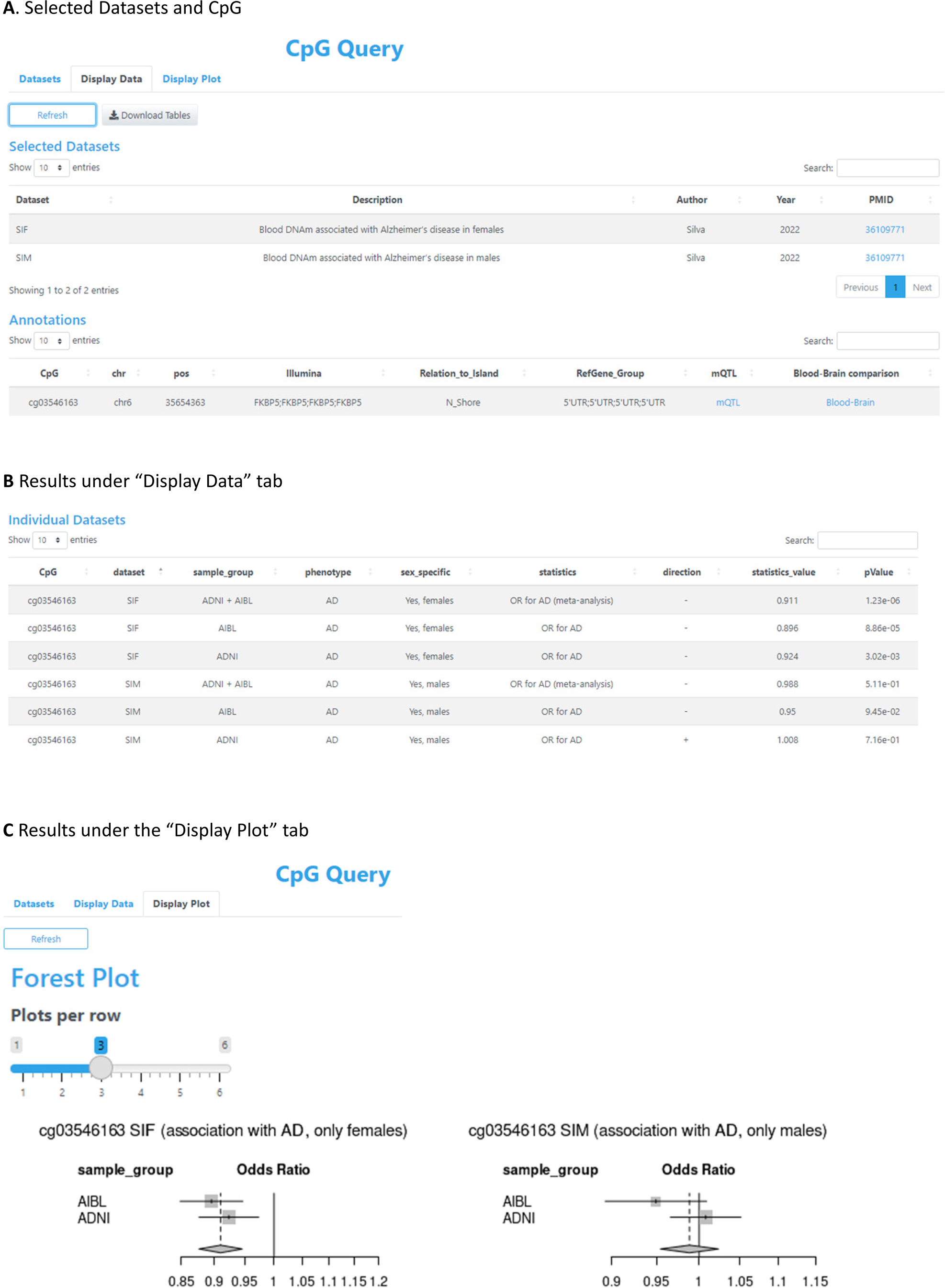
The CpG Query Tool can be used to explore details of DNA methylation at female-specific CpG cg03546163 associated with AD.

## References

1. Rajan KB, Weuve J, Barnes LL, McAninch EA, Wilson RS, Evans DA: Population estimate of people with clinical Alzheimer’s disease and mild cognitive impairment in the United States (2020-2060). Alzheimers Dement 2021, 17:1966–1975.

2. Hurd MD, Martorell P, Delavande A, Mullen KJ, Langa KM: Monetary costs of dementia in the United States. N Engl J Med 2013, 368:1326–1334.

3. Hudomiet P, Hurd MD, Rohwedder S: Dementia Prevalence in the United States in 2000 and 2012: Estimates Based on a Nationally Representative Study. J Gerontol B Psychol Sci Soc Sci 2018, 73:S10–S19.

4. Plassman BL, Langa KM, Fisher GG, Heeringa SG, Weir DR, Ofstedal MB, Burke JR, Hurd MD, Potter GG, Rodgers WL, et al: Prevalence of dementia in the United States: the aging, demographics, and memory study. Neuroepidemiology 2007, 29:125–132.

5. Ferretti MT, Iulita MF, Cavedo E, Chiesa PA, Schumacher Dimech A, Santuccione Chadha A, Baracchi F, Girouard H, Misoch S, Giacobini E, et al: Sex differences in Alzheimer disease - the gateway to precision medicine. Nat Rev Neurol 2018, 14:457–469.

6. Mazure CM, Swendsen J: Sex differences in Alzheimer’s disease and other dementias. Lancet Neurol 2016, 15:451–452.

7. Nebel RA, Aggarwal NT, Barnes LL, Gallagher A, Goldstein JM, Kantarci K, Mallampalli MP, Mormino EC, Scott L, Yu WH, et al: Understanding the impact of sex and gender in Alzheimer’s disease: A call to action. Alzheimers Dement 2018, 14:1171–1183.

8. Hampel H, Vergallo A, Giorgi FS, Kim SH, Depypere H, Graziani M, Saidi A, Nistico R, Lista S, Alzheimer Precision Medicine I: Precision medicine and drug development in Alzheimer’s disease: the importance of sexual dimorphism and patient stratification. Front Neuroendocrinol 2018, 50:31–51.

9. Lin KA, Doraiswamy PM: When Mars Versus Venus is Not a Cliche: Gender Differences in the Neurobiology of Alzheimer’s Disease. Front Neurol 2014, 5:288.

10. Hebert LE, Weuve J, Scherr PA, Evans DA: Alzheimer disease in the United States (2010-2050) estimated using the 2010 census. Neurology 2013, 80:1778–1783.

11. Koran MEI, Wagener M, Hohman TJ, Alzheimer’s Neuroimaging I: Sex differences in the association between AD biomarkers and cognitive decline. Brain Imaging Behav 2017, 11:205–213.

12. Tschanz JT, Corcoran CD, Schwartz S, Treiber K, Green RC, Norton MC, Mielke MM, Piercy K, Steinberg M, Rabins PV, et al: Progression of cognitive, functional, and neuropsychiatric symptom domains in a population cohort with Alzheimer dementia: the Cache County Dementia Progression study. Am J Geriatr Psychiatry 2011, 19:532–542.

13. Portela A, Esteller M: Epigenetic modifications and human disease. Nat Biotechnol 2010, 28:1057–1068.

14. Bell CG, Xia Y, Yuan W, Gao F, Ward K, Roos L, Mangino M, Hysi PG, Bell J, Wang J, Spector TD: Novel regional age-associated DNA methylation changes within human common disease-associated loci. Genome Biol 2016, 17:193.

15. Day K, Waite LL, Thalacker-Mercer A, West A, Bamman MM, Brooks JD, Myers RM, Absher D: Differential DNA methylation with age displays both common and dynamic features across human tissues that are influenced by CpG landscape. Genome Biol 2013, 14:R102.

16. Johansson A, Enroth S, Gyllensten U: Continuous Aging of the Human DNA Methylome Throughout the Human Lifespan. PLoS One 2013, 8:e67378.

17. Jones MJ, Goodman SJ, Kobor MS: DNA methylation and healthy human aging. Aging Cell 2015, 14:924–932.

18. Lord J, Cruchaga C: The epigenetic landscape of Alzheimer’s disease. Nat Neurosci 2014, 17:1138–1140.

19. Lunnon K, Smith R, Hannon E, De Jager PL, Srivastava G, Volta M, Troakes C, Al-Sarraj S, Burrage J, Macdonald R, et al: Methylomic profiling implicates cortical deregulation of ANK1 in Alzheimer’s disease. Nat Neurosci 2014, 17:1164–1170.

20. Stoccoro A, Coppede F: Role of epigenetics in Alzheimer’s disease pathogenesis. Neurodegener Dis Manag 2018, 8:181–193.

21. Fenoglio C, Scarpini E, Serpente M, Galimberti D: Role of Genetics and Epigenetics in the Pathogenesis of Alzheimer’s Disease and Frontotemporal Dementia. J Alzheimers Dis 2018, 62:913–932.

22. Roubroeks JAY, Smith RG, van den Hove DLA, Lunnon K: Epigenetics and DNA methylomic profiling in Alzheimer’s disease and other neurodegenerative diseases. J Neurochem 2017, 143:158–170.

23. Bacalini MG, Boattini A, Gentilini D, Giampieri E, Pirazzini C, Giuliani C, Fontanesi E, Remondini D, Capri M, Del Rio A, et al: A meta-analysis on age-associated changes in blood DNA methylation: results from an original analysis pipeline for Infinium 450k data. Aging (Albany NY) 2015, 7:97–109.

24. Reynolds LM, Taylor JR, Ding J, Lohman K, Johnson C, Siscovick D, Burke G, Post W, Shea S, Jacobs DR, Jr., et al: Age-related variations in the methylome associated with gene expression in human monocytes and T cells. Nat Commun 2014, 5:5366.

25. Chen BH, Marioni RE, Colicino E, Peters MJ, Ward-Caviness CK, Tsai PC, Roetker NS, Just AC, Demerath EW, Guan W, et al: DNA methylation-based measures of biological age: meta-analysis predicting time to death. Aging (Albany NY) 2016, 8:1844–1865.

26. Horvath S, Raj K: DNA methylation-based biomarkers and the epigenetic clock theory of ageing. Nat Rev Genet 2018, 19:371–384.

27. Zhang L, Silva TC, Young JI, Gomez L, Schmidt MA, Hamilton-Nelson KL, Kunkle BW, Chen X, Martin ER, Wang L: Epigenome-wide meta-analysis of DNA methylation differences in prefrontal cortex implicates the immune processes in Alzheimer’s disease. Nat Commun 2020, 11:6114.

28. Zhang L, Young JI, Gomez L, Silva TC, Schmidt MA, Cai J, Chen X, Martin ER, Wang L: Sex-specific DNA methylation differences in Alzheimer’s disease pathology. Acta Neuropathol Commun 2021, 9:77.

29. T CS, Zhang W, Young JI, Gomez L, Schmidt MA, Varma A, Chen XS, Martin ER, Wang L: Distinct sex-specific DNA methylation differences in Alzheimer’s disease. Alzheimers Res Ther 2022, 14:133.

30. T CS, Young JI, Zhang L, Gomez L, Schmidt MA, Varma A, Chen XS, Martin ER, Wang L: Cross-tissue analysis of blood and brain epigenome-wide association studies in Alzheimer’s disease. Nat Commun 2022, 13:4852.

31. De Jager PL, Srivastava G, Lunnon K, Burgess J, Schalkwyk LC, Yu L, Eaton ML, Keenan BT, Ernst J, McCabe C, et al: Alzheimer’s disease: early alterations in brain DNA methylation at ANK1, BIN1, RHBDF2 and other loci. Nat Neurosci 2014, 17:1156–1163.

32. Smith RG, Hannon E, De Jager PL, Chibnik L, Lott SJ, Condliffe D, Smith AR, Haroutunian V, Troakes C, Al-Sarraj S, et al: Elevated DNA methylation across a 48-kb region spanning the HOXA gene cluster is associated with Alzheimer’s disease neuropathology. Alzheimers Dement 2018, 14:1580–1588.

33. Smith RG, Pishva E, Shireby G, Smith AR, Roubroeks JAY, Hannon E, Wheildon G, Mastroeni D, Gasparoni G, Riemenschneider M, et al: A meta-analysis of epigenome-wide association studies in Alzheimer’s disease highlights novel differentially methylated loci across cortex. Nat Commun 2021, 12:3517.

34. Fransquet PD, Lacaze P, Saffery R, McNeil J, Woods R, Ryan J: Blood DNA methylation as a potential biomarker of dementia: A systematic review. Alzheimers Dement 2018, 14:81–103.

35. Kobayashi N, Shinagawa S, Niimura H, Kida H, Nagata T, Tagai K, Shimada K, Oka N, Shikimoto R, Noda Y, et al: Increased blood COASY DNA methylation levels a potential biomarker for early pathology of Alzheimer’s disease. Sci Rep 2020, 10:12217.

36. Roubroeks JAY, Smith AR, Smith RG, Pishva E, Ibrahim Z, Sattlecker M, Hannon EJ, Kloszewska I, Mecocci P, Soininen H, et al: An epigenome-wide association study of Alzheimer’s disease blood highlights robust DNA hypermethylation in the HOXB6 gene. Neurobiol Aging 2020, 95:26–45.

37. Fransquet PD, Lacaze P, Saffery R, Phung J, Parker E, Shah RC, Murray A, Woods RL, Ryan J: DNA methylation analysis of candidate genes associated with dementia in peripheral blood. Epigenomics 2020, 12:2109–2123.

38. Madrid A, Hogan KJ, Papale LA, Clark LR, Asthana S, Johnson SC, Alisch RS: DNA Hypomethylation in Blood Links B3GALT4 and ZADH2 to Alzheimer’s Disease. J Alzheimers Dis 2018, 66:927–934.

39. Mitsumori R, Sakaguchi K, Shigemizu D, Mori T, Akiyama S, Ozaki K, Niida S, Shimoda N: Lower DNA methylation levels in CpG island shores of CR1, CLU, and PICALM in the blood of Japanese Alzheimer’s disease patients. PLoS One 2020, 15:e0239196.

40. Vasanthakumar A, Davis JW, Idler K, Waring JF, Asque E, Riley-Gillis B, Grosskurth S, Srivastava G, Kim S, Nho K, et al: Harnessing peripheral DNA methylation differences in the Alzheimer’s Disease Neuroimaging Initiative (ADNI) to reveal novel biomarkers of disease. Clin Epigenetics 2020, 12:84.

41. Nabais MF, Laws SM, Lin T, Vallerga CL, Armstrong NJ, Blair IP, Kwok JB, Mather KA, Mellick GD, Sachdev PS, et al: Meta-analysis of genome-wide DNA methylation identifies shared associations across neurodegenerative disorders. Genome Biol 2021, 22:90.

42. Mikeska T, Craig JM: DNA methylation biomarkers: cancer and beyond. Genes (Basel) 2014, 5:821–864.

43. Tacutu R, Craig T, Budovsky A, Wuttke D, Lehmann G, Taranukha D, Costa J, Fraifeld VE, de Magalhaes JP: Human Ageing Genomic Resources: integrated databases and tools for the biology and genetics of ageing. Nucleic Acids Res 2013, 41:D1027–1033.

44. Li M, Zou D, Li Z, Gao R, Sang J, Zhang Y, Li R, Xia L, Zhang T, Niu G, et al: EWAS Atlas: a curated knowledgebase of epigenome-wide association studies. Nucleic Acids Res 2019, 47:D983–D988.

45. Battram T, Yousefi P, Crawford G, Prince C, Sheikhali Babaei M, Sharp G, Hatcher C, Vega-Salas MJ, Khodabakhsh S, Whitehurst O, et al: The EWAS Catalog: a database of epigenome-wide association studies. Wellcome Open Res 2022, 7:41.

46. Chang W, Cheng J, Allaire J, Sievert C, Schloerke B, Xie Y, Allen J, McPherson J, Dipert A, Borges B: shiny: Web Application Framework for R. R package version 174 2022:https://CRAN.R-project.org/package=shiny.

47. Attali D: shinyjs: Easily Improve the User Experience of Your Shiny Apps in Seconds. R pakcage version 210 2021:https://CRAN.R-project.org/package=shinyjs.

48. Perrier V, Meyer F, Granjon D: shinyWidgets: Custom Inputs Widgets for Shiny R package version 076 2023:https://CRAN.R-project.org/package=shinyWidgets.

49. Hahne F, Ivanek R: Visualizing Genomic Data Using Gviz and Bioconductor. Methods Mol Biol 2016, 1418:335–351.

50. Zhou X, Li D, Zhang B, Lowdon RF, Rockweiler NB, Sears RL, Madden PA, Smirnov I, Costello JF, Wang T: Epigenomic annotation of genetic variants using the Roadmap Epigenome Browser. Nat Biotechnol 2015, 33:345–346.

51. Min JL, Hemani G, Hannon E, Dekkers KF, Castillo-Fernandez J, Luijk R, Carnero-Montoro E, Lawson DJ, Burrows K, Suderman M, et al: Genomic and phenotypic insights from an atlas of genetic effects on DNA methylation. Nat Genet 2021, 53:1311–1321.

52. Hannon E, Lunnon K, Schalkwyk L, Mill J: Interindividual methylomic variation across blood, cortex, and cerebellum: implications for epigenetic studies of neurological and neuropsychiatric phenotypes. Epigenetics 2015, 10:1024–1032.

53. Horvath S: DNA methylation age of human tissues and cell types. Genome Biol 2013, 14:R115.

54. Sugden K, Caspi A, Elliott ML, Bourassa KJ, Chamarti K, Corcoran DL, Hariri AR, Houts RM, Kothari M, Kritchevsky S, et al: Association of Pace of Aging Measured by Blood-Based DNA Methylation With Age-Related Cognitive Impairment and Dementia. Neurology 2022, 99:e1402–e1413.

55. Kennedy BK, Berger SL, Brunet A, Campisi J, Cuervo AM, Epel ES, Franceschi C, Lithgow GJ, Morimoto RI, Pessin JE, et al: Geroscience: linking aging to chronic disease. Cell 2014, 159:709–713.

56. McKinney BC, Sibille E: The age-by-disease interaction hypothesis of late-life depression. Am J Geriatr Psychiatry 2013, 21:418–432.

57. McKinney BC, Lin CW, Rahman T, Oh H, Lewis DA, Tseng G, Sibille E: DNA methylation in the human frontal cortex reveals a putative mechanism for age-by-disease interactions. Transl Psychiatry 2019, 9:39.

58. Nativio R, Donahue G, Berson A, Lan Y, Amlie-Wolf A, Tuzer F, Toledo JB, Gosai SJ, Gregory BD, Torres C, et al: Dysregulation of the epigenetic landscape of normal aging in Alzheimer’s disease. Nat Neurosci 2018, 21:497–505.

59. Nativio R, Lan Y, Donahue G, Sidoli S, Berson A, Srinivasan AR, Shcherbakova O, Amlie-Wolf A, Nie J, Cui X, et al: An integrated multi-omics approach identifies epigenetic alterations associated with Alzheimer’s disease. Nat Genet 2020, 52:1024–1035.

60. McCartney DL, Zhang F, Hillary RF, Zhang Q, Stevenson AJ, Walker RM, Bermingham ML, Boutin T, Morris SW, Campbell A, et al: An epigenome-wide association study of sex-specific chronological ageing. Genome Med 2019, 12:1.

61. Rattan SI: Synthesis, modifications, and turnover of proteins during aging. Exp Gerontol 1996, 31:33–47.

62. Anisimova AS, Alexandrov AI, Makarova NE, Gladyshev VN, Dmitriev SE: Protein synthesis and quality control in aging. Aging (Albany NY) 2018, 10:4269–4288.

63. Gonskikh Y, Polacek N: Alterations of the translation apparatus during aging and stress response. Mech Ageing Dev 2017, 168:30–36.

64. Lee AG, Capanzana R, Brockhurst J, Cheng MY, Buckmaster CL, Absher D, Schatzberg AF, Lyons DM: Learning to cope with stress modulates anterior cingulate cortex stargazin expression in monkeys and mice. Neurobiol Learn Mem 2016, 131:95–100.

65. Huganir RL, Nicoll RA: AMPARs and synaptic plasticity: the last 25 years. Neuron 2013, 80:704–717.

66. Beneyto M, Meador-Woodruff JH: Lamina-specific abnormalities of AMPA receptor trafficking and signaling molecule transcripts in the prefrontal cortex in schizophrenia. Synapse 2006, 60:585–598.

67. Quarato V, D’’Antona S, Battista P, Zupo R, Sardone R, Castiglioni I, Porro D, Frasca M, Cava C: Transcriptional Profiling of Hippocampus Identifies Network Alterations in Alzheimer’s Disease. Applied Sciences 2022, 12:5035.

68. Pelucchi S, Gardoni F, Di Luca M, Marcello E: Synaptic dysfunction in early phases of Alzheimer’s Disease. Handb Clin Neurol 2022, 184:417–438.

69. Esiri MM, Chance SA: Cognitive reserve, cortical plasticity and resistance to Alzheimer’s disease. Alzheimers Res Ther 2012, 4:7.

70. Martorana A, Assogna M, V DEL, Motta C, Bonomi CG, Bernocchi F, MG DID, Koch G: Cognitive reserve and Alzheimer’s biological continuum: clues for prediction and prevention of dementia. Minerva Med 2021, 112:441–447.

71. Binder EB, Salyakina D, Lichtner P, Wochnik GM, Ising M, Putz B, Papiol S, Seaman S, Lucae S, Kohli MA, et al: Polymorphisms in FKBP5 are associated with increased recurrence of depressive episodes and rapid response to antidepressant treatment. Nat Genet 2004, 36:1319–1325.

72. Hernandez-Diaz Y, Gonzalez-Castro TB, Tovilla-Zarate CA, Juarez-Rojop IE, Lopez-Narvaez ML, Perez-Hernandez N, Rodriguez-Perez JM, Genis-Mendoza AD: Association between FKBP5 polymorphisms and depressive disorders or suicidal behavior: A systematic review and meta-analysis study. Psychiatry Res 2019, 271:658–668.

73. Abate KH: Gender disparity in prevalence of depression among patient population: a systematic review. Ethiop J Health Sci 2013, 23:283–288.

74. Figueroa PBS, Ferreira AFF, Britto LR, Doussoulin AP, Torrao ADS: Association between thyroid function and Alzheimer’s disease: A systematic review. Metab Brain Dis 2021, 36:1523–1543.

75. Salehipour A, Dolatshahi M, Haghshomar M, Amin J: The Role of Thyroid Dysfunction in Alzheimer’s Disease: A Systematic Review and Meta-Analysis. J Prev Alzheimers Dis 2023, 10:276–286.

76. Aging Atlas C: Aging Atlas: a multi-omics database for aging biology. Nucleic Acids Res 2021, 49:D825–D830.

